# Serum S100A8/S100A9 is associated with increased risk of brain metastasis in patients with inflammatory breast cancer

**DOI:** 10.64898/2026.01.21.26344294

**Authors:** Emilly S Villodre, Juhee Song, Xiaoding Hu, Kristen Gomez, Evan N. Cohen, James M. Reuben, Azadeh Nasrazadani, Bora Lim, The MDACC Inflammatory Breast Cancer Team, Debu Tripathy, Wendy A. Woodward, Savitri Krishnamurthy, Bisrat G Debeb

## Abstract

**Background:** Inflammatory breast cancer (IBC) is a rare and highly aggressive form of breast cancer with an increased propensity to metastasize to distant organs including the brain. Higher serum levels of the calcium-binding proteins S100A8/A9, particularly of S100A9, have emerged as a clinically and biologically significant factor in aggressive breast cancers that are associated with poorer prognosis, tumor progression, and resistance to therapy. However, its contribution in IBC specifically remains undefined. Here, we investigated whether serum levels of S100A8/A9 predict outcomes in patients with IBC.

**Methods:** Serum S100A8/A9 levels were measured in a cohort of 304 IBC patients using ELISA assay. S100A8/A9 levels were categorized by their third quartile value (S100A8/A9-low ≤ 3^rd^ quartile; S100A8/A9-high > 3^rd^ quartile). Overall survival (OS) and breast cancer-specific survival (BCSS) were analyzed with Kaplan-Meier curves, log-rank tests, and Cox proportional hazard regression models. The cumulative incidence of any metastases and the cumulative incidence of brain metastases were analyzed using Aalen-Johansen method, Gray test, and Fine-Gray models.

**Results:** The median follow-up time was 64 months. Forty-six percent of patients had estrogen receptor (ER)-negative tumors, 61.3% were stage III-IV, 77% high grade, 16.8% received adjuvant chemotherapy and 53.6% received adjuvant radiation. On univariate analysis, S100A8/A9 levels, disease stage, ER status, PR status, HER2 status, adjuvant chemotherapy, and adjuvant radiation therapy were significantly associated with OS and BCSS. Patients with high S100A8/A9 serum levels had poor OS (*P*=0.01) and BCSS (*P*=0.007) and had a higher risk of developing brain metastasis (*P*=0.01) but not other metastasis. On multivariate analysis, high S100A8/A9 serum levels were independently associated with reduced OS (hazard ratio [HR]=1.7, 95% CI 1.1 to 2.6, *P*=0.01), reduced BCSS (HR=1.8, 95% CI 1.2 to 2.8, *P*=0.006), and increased cumulative incidence of developing brain metastasis (subdistribution hazard ratio (sHR)=1.8, 95% CI 1.1 to 3.0, *P*=0.03).

**Conclusions:** In patients with IBC, high serum levels of S100A8/A9 are an independent prognostic factor for brain metastasis and poor clinical outcomes. These findings support the potential of S100A8/A9 as predictive biomarker for identifying increased risk of brain metastasis and unfavorable prognosis in patients with IBC.

## Introduction

Inflammatory breast cancer (IBC) represents one of the most aggressive subtypes of breast cancer. Although it is uncommon, contributing just 1% to 4% of newly diagnosed breast cancer cases, it accounts for an outsized 10% of breast cancer-related deaths in the United States^1,2^. IBC is marked by a distinct clinical profile, including rapid tumor growth and early spread. Nearly all patients diagnosed with IBC show lymph node involvement, and over one-third have distant metastases at the time of diagnosis^3,4^. Despite treatment strategies that integrate systemic chemotherapy, surgery, and radiation, outcomes for IBC patients are significantly poorer than for those with non-inflammatory breast cancer, with 5-year overall survival (OS) rates of 40% compared with 63%^5-7^. Significant efforts have been undertaken to discover molecular markers and therapeutic targets specific to IBC, and several have been proposed, including EGFR, E-cadherin, eIFG4I, RhoC, and TIG1/AXL^8-12^. However, no definitive molecular target or signature unique to IBC has been identified and validated yet, and effective targeted treatment options remain scarce.

S100A8 and S100A9 are calcium-binding proteins primarily expressed by myeloid cells, which form heterodimers known as calprotectin that regulate inflammation and immune cell recruitment^13^. In cancer, including breast cancer, elevated S100A8/A9 levels contribute to tumor proliferation, invasion, and metastasis by activating receptors like TLR4 and RAGE, thereby promoting pro-inflammatory signaling and immune evasion^13-15^. These proteins also facilitate metastasis by recruiting immunosuppressive cells such as myeloid-derived suppressor cells to pre-metastatic niches, particularly in the lungs and brain^13,14,16-18^. Recent studies elegantly demonstrated a critical role for S100A9 in brain relapse and radioresistance in brain metastasis mouse models^19,20^.

Clinically, high S100A9 levels have been found to be a prognostic indicator for disease progression and lower overall survival (OS) as well as to predict therapeutic resistance, particularly in the context of immunotherapies^20-26^. S100A8/A9 has emerged as a critical mediator in triple-negative breast cancer (TNBC), with relevance as both a biomarker and a potential therapeutic target. Elevated serum S100A8/A9 levels have been associated with more aggressive disease subtypes and poor patient outcomes, indicating its prognostic value^27^. Mechanistic studies further reveal that S100A8/A9 promotes tumor progression by modulating the tumor microenvironment and driving pro-tumorigenic inflammation, supporting its role as a functional contributor to TNBC pathophysiology^28^. Importantly, preclinical models demonstrate that targeting S100A8/A9, either directly or indirectly, can enhance therapeutic responses, including in combination with immune checkpoint blockade or kinase inhibitors, highlighting its potential as a predictive and pharmacodynamic biomarker^29^. Together, these findings support the investigation of serum S100A8/A9 not only as a prognostic indicator but also as a therapeutic-relevant biomarker in aggressive breast cancer. Despite these advances, the contribution of S100A8/A9 to the biology and clinical behavior of inflammatory breast cancer remains largely unexplored, underscoring a critical gap in our understanding of this highly aggressive subtype.

In this study, we investigated S100A8/A9 levels in serum samples from patients with IBC and demonstrated that elevated S100A8/A9 is an independent predictor of poorer OS, breast cancer–specific survival (BCSS), and development of brain metastases.

## Results

### S100A8/A9 is associated with poor outcome in IBC patients

To assess potential associations between S100A8/A9 serum levels and outcomes in IBC, we used an enzyme-linked immunosorbent assay of serum samples from a retrospective cohort of 304 patients with IBC. The median age of these patients was 52 years (range 23–81 years). The median follow-up time was 64 months. Forty-six percent of patients had estrogen receptor (ER)-negative tumors, 61.3% had stage III-IV disease, 77% had high-grade disease, 16.8% received adjuvant chemotherapy, and 53.6% received adjuvant radiation. Table 1 summarizes patient characteristics for the entire group.

**Table 1.**
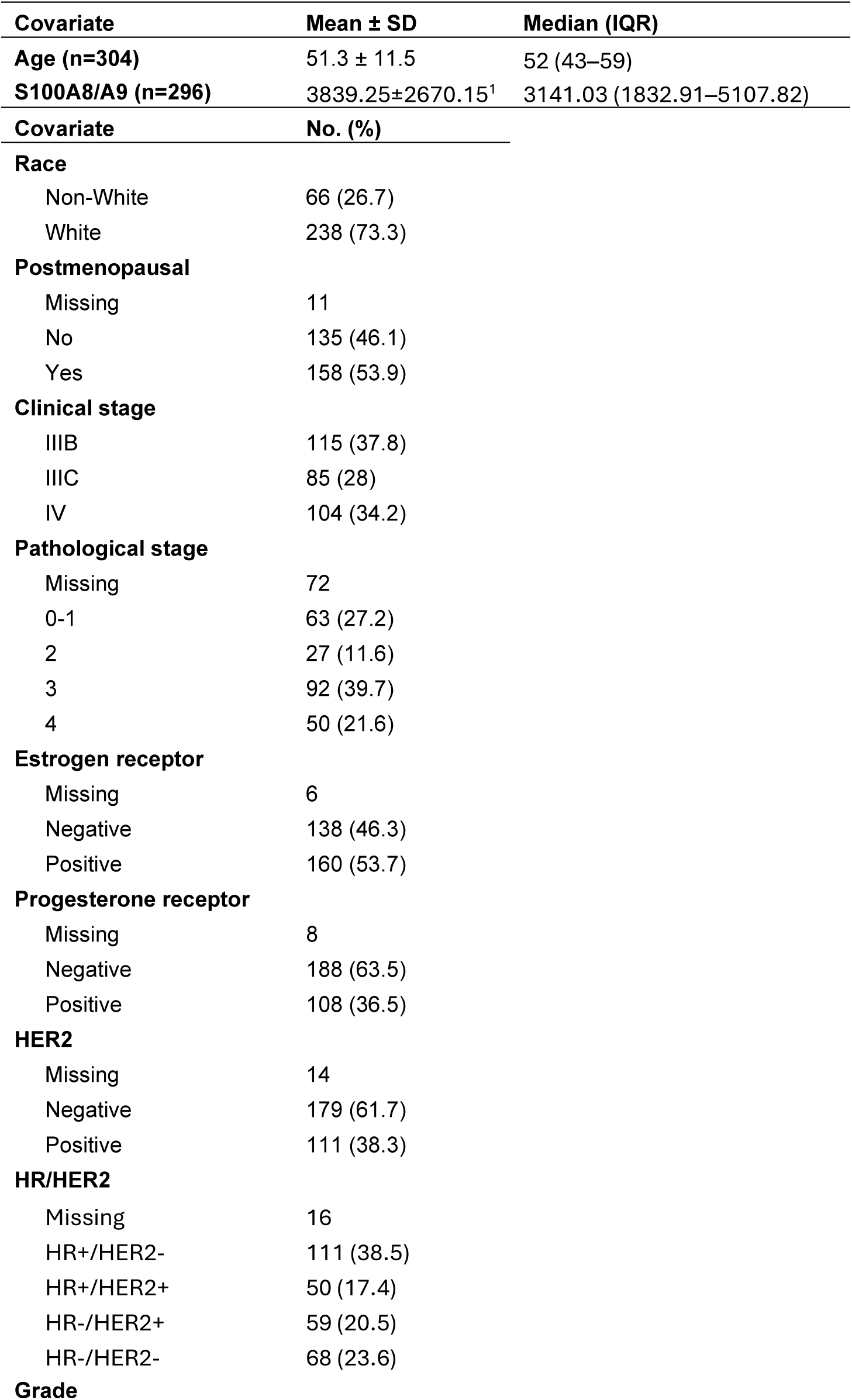
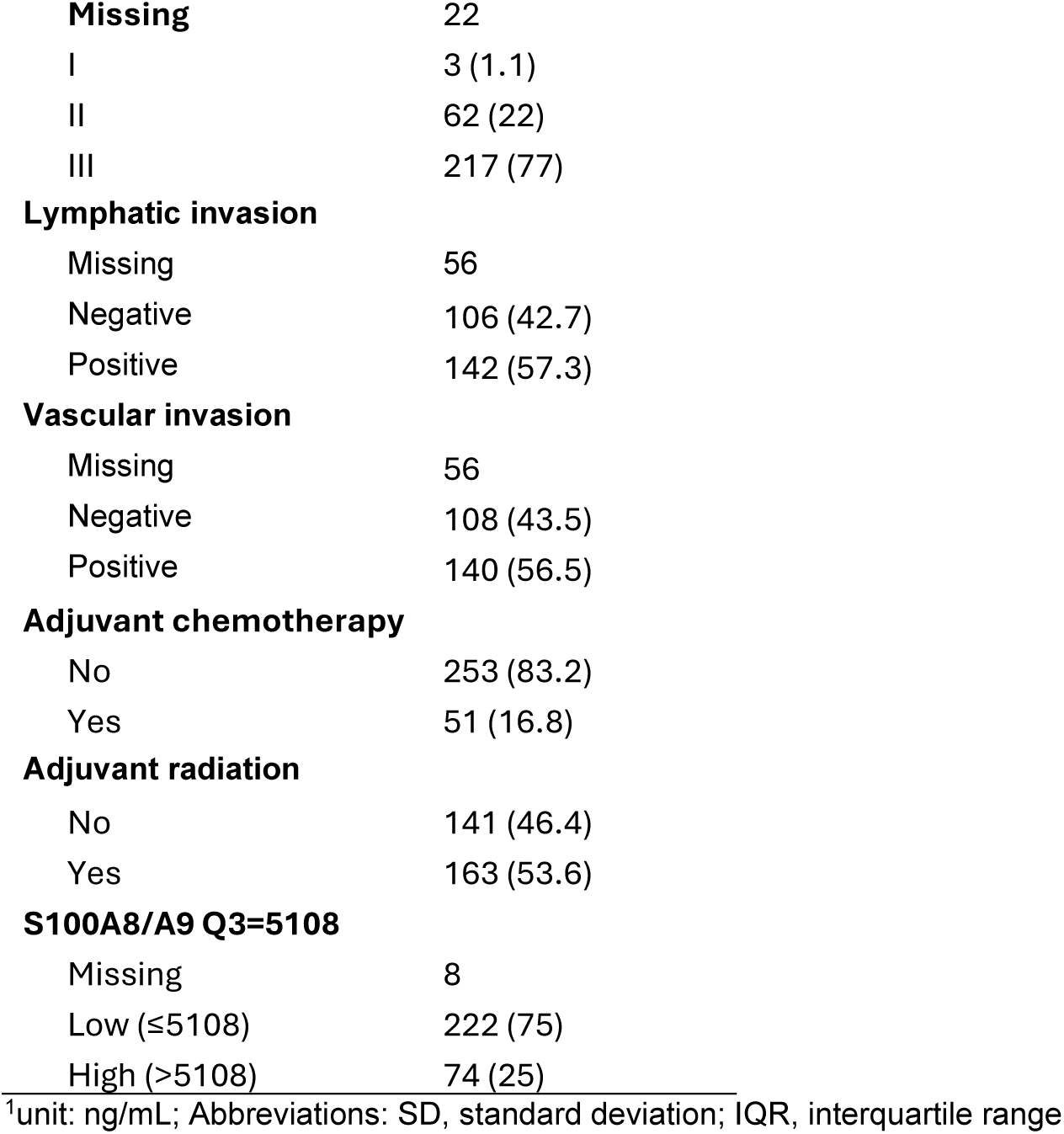
Clinical and pathologic characteristics of tumor samples from patients with IBC.

| **Covariate** | **Mean ± SD** | **Median (IQR)** |
| --- | --- | --- |
| **Age (n=304)** | 51.3 ± 11.5 | 52 (43–59) |
| **S100A8/A9 (n=296)** | 3839.25±2670.15<sup>1</sup> | 3141.03 (1832.91–5107.82) |
| **Covariate** | **No. (%)** |  |
| **Race** |  |  |
| Non-White | 66 (26.7) |  |
| White | 238 (73.3) |  |
| **Postmenopausal** |  |  |
| Missing | 11 |  |
| No | 135 (46.1) |  |
| Yes | 158 (53.9) |  |
| **Clinical stage** |  |  |
| IIIB | 115 (37.8) |  |
| IIIC | 85 (28) |  |
| IV | 104 (34.2) |  |
| **Pathological stage** |  |  |
| Missing | 72 |  |
| 0-1 | 63 (27.2) |  |
| 2 | 27 (11.6) |  |
| 3 | 92 (39.7) |  |
| 4 | 50 (21.6) |  |
| **Estrogen receptor** |  |  |
| Missing | 6 |  |
| Negative | 138 (46.3) |  |
| Positive | 160 (53.7) |  |
| **Progesterone receptor** |  |  |
| Missing | 8 |  |
| Negative | 188 (63.5) |  |
| Positive | 108 (36.5) |  |
| **HER2** |  |  |
| Missing | 14 |  |
| Negative | 179 (61.7) |  |
| Positive | 111 (38.3) |  |
| **HR/HER2** |  |  |
| Missing | 16 |  |
| HR+/HER2- | 111 (38.5) |  |
| HR+/HER2+ | 50 (17.4) |  |
| HR-/HER2+ | 59 (20.5) |  |
| HR-/HER2- | 68 (23.6) |  |
| **Grade** |  |  |

|  |  |
| --- | --- |
| **Missing** | 22 |
| I | 3 (1.1) |
| II | 62 (22) |
| III | 217 (77) |
| **Lymphatic invasion** |  |
| Missing | 56 |
| Negative | 106 (42.7) |
| Positive | 142 (57.3) |
| **Vascular invasion** |  |
| Missing | 56 |
| Negative | 108 (43.5) |
| Positive | 140 (56.5) |
| **Adjuvant chemotherapy** |  |
| No | 253 (83.2) |
| Yes | 51 (16.8) |
| **Adjuvant radiation** |  |
| No | 141 (46.4) |
| Yes | 163 (53.6) |
| **S100A8/A9 Q3=5108** |  |
| Missing | 8 |
| Low ($\leq 5108$) | 222 (75) |
| High ($> 5108$) | 74 (25) |

<sup>†</sup>unit: ng/mL; Abbreviations: SD, standard deviation; IQR, interquartile range

Univariate analysis (Table 2) identified significant associations between OS and S100A8/A9 serum levels (hazard ratio [HR] =1.67, *P*=0.0120), estrogen receptor (ER) status (HR=0.67, *P*=0.0280), HER2 status (HR=0.28, *P*<0.0001) and receipt of adjuvant radiation therapy (HR=0.37, *P*<0.0001). These variables were similarly associated with BCSS (Table 2). In addition, analysis of metastasis-related outcomes revealed that S100A8/A9 expression was significantly associated with the development of brain metastases (subdistribution hazard ratio [sHR]=1.88, *P*=0.018), but not with the development with metastases at other sites (*P*=0.348) (Table 3).

**Table 2.** Univariate Cox regression analysis on overall survival and breast cancer specific survival among patients with IBC.

| Covariate | Overall survival |  |  | Breast cancer specific survival |  |  |
| --- | --- | --- | --- | --- | --- | --- |
|  | HR | 95% CI | P Value | HR | 95% CI | P Value |
| **Age** |  |  |  |  |  |  |
| 1 Unit Change | 1.004 | 0.988–1.019 | 0.6536 | 1.000 | 0.984–1.016 | 0.9868 |
| **S100A9** |  |  |  |  |  |  |
| Low | 1.000 |  |  | 1.000 |  |  |
| High | 1.667 | 1.119–2.483 | **0.0120** | 1.755 | 1.168–2.639 | **0.0068** |
| **Race** |  |  |  |  |  |  |
| Non-black | 1.000 |  |  | 1.000 |  |  |
| Black | 1.793 | 1.075–2.991 | **0.0253** | 1.921 | 1.149–3.212 | **0.0128** |
| **Clinical Stage** |  |  |  |  |  |  |
| IIIb | 1.000 |  |  | 1.000 |  |  |
| IIIc | 1.947 | 1.125–3.092 | **0.0048** | 1.940 | 1.193–3.156 | **0.0076** |
| IV | 2.406 | 1.552–3.730 | **<0.0001** | 2.661 | 1.693–3.156 | **<0.0001** |
| **Pathological Stage** |  |  |  |  |  |  |
| 0-1 | 1.000 |  |  | 1.000 |  |  |
| 2 | 1.976 | 0.818–4.773 | **0.1300** | 2.145 | 0.827–5.563 | 0.1167 |
| 3 | 3.097 | 1.590–6.031 | **0.0009** | 3.430 | 1.654–7.116 | **0.0009** |
| 4 | 2.769 | 1.350–5.682 | **0.0055** | 3.386 | 1.566–7.319 | **0.0019** |
| **Estrogen receptor** |  |  |  |  |  |  |
| Negative | 1.000 |  |  | 1.000 |  |  |
| Positive | 0.673 | 0.473–0.958 | **0.0280** | 0.656 | 0.456–0.943 | **0.0230** |
| **Progesterone receptor** |  |  |  |  |  |  |
| Negative | 1.000 |  |  | 1.000 |  |  |
| Positive | 0.456 | 0.302–0.689 | **0.0002** | 0.471 | 0.309–0.718 | **0.0005** |
| **Hormone receptor** |  |  |  |  |  |  |
| Negative | 1.000 |  |  | 1.000 |  |  |
| Positive | 0.683 | 0.480–0.973 | **0.0348** | 0.669 | 0.465–0.963 | **0.0305** |
| **HER2** |  |  |  |  |  |  |
| Negative | 1.000 |  |  | 1.000 |  |  |
| Positive | 0.277 | 0.174–0.441 | **<0.0001** | 0.281 | 0.174–0.453 | **<0.0001** |
| **Hormone receptor / HER2** |  |  |  |  |  |  |
| HR+/HER2- | 1.000 |  |  | 1.000 |  |  |
| HR+/HER2+ | 0.392 | 0.198–0.775 | **0.0071** | 0.376 | 0.183–0.770 | **0.0075** |
| HR-/HER2+ | 0.383 | 0.203–0.723 | **0.0031** | 0.407 | 0.214–0.772 | **0.0059** |
| HR-/HER2- | 2.137 | 1.427–3.200 | **0.0002** | 2.148 | 1.416–3.260 | **0.0003** |
| **Grade** |  |  |  |  |  |  |
| I-II | 1.000 |  |  | 1.000 |  |  |
| III | 1.089 | 0.700–1.695 | 0.7058 | 1.124 | 0.709–1.780 | 0.6194 |
| **Lymphatic invasion<sup>1</sup>** |  |  |  |  |  |  |
| Negative | 1.000 |  |  | 1.000 |  |  |
| Positive | 1.945 | 1.266–2.989 | **0.0024** | 2.152 | 1.367–3.388 | **0.0009** |
| **Vascular invasion<sup>1</sup>** |  |  |  |  |  |  |
| Negative | 1.000 |  |  | 1.000 |  |  |
| Positive | 2.027 | 1.319–3.112 | **0.0013** | 2.241 | 1.424–3.528 | **0.0005** |
| **Adjuvant chemotherapy** |  |  |  |  |  |  |
| No | 1.000 |  |  | 1.000 |  |  |
| Yes | 0.451 | 0.262–0.779 | **0.0043** | 0.442 | 0.251–0.776 | **0.0045** |
| Yes | 1.770 | 0.561–5.587 | 0.3305 | 1.855 | 0.587–5.864 | 0.2925 |
| **Adjuvant radiation** |  |  |  |  |  |  |
| No | 1.000 |  |  | 1.000 |  |  |
| Yes | 0.368 | 0.257–0.525 | **<0.0001** | 0.339 | 0.234–0.491 | **<0.0001** |

<sup>1</sup>These covariates were not considered in multivariable models due to too many missing measurements.

**Table 3.** Univariate Fine-Gray regression analysis on development of any metastases (n=187) or brain metastases (n=62) among patients with IBC.

| Covariate | Overall survival |  |  | Breast cancer specific survival |  |  |
| --- | --- | --- | --- | --- | --- | --- |
|  | HR | 95% CI | P Value | HR | 95% CI | P Value |
| **Age** |  |  |  |  |  |  |
| 1 Unit Change | 0.996 | 0.984–1.008 | 0.5281 | 0.976 | 0.955–0.998 | **0.0299** |
| **S100A9** |  |  |  |  |  |  |
| Low | 1.000 |  |  | 1.000 |  |  |
| High | 1.162 | 0.849–1.589 | 0.3480 | 1.883 | 0.113–3.185 | **0.0184** |
| **Race** |  |  |  |  |  |  |
| Non-black | 1.000 |  |  | 1.000 |  |  |
| Black | 1.626 | 1.138–2.324 | **0.0077** | 1.010 | 0.441–2.316 | 0.9804 |
| **Clinical Stage** |  |  |  |  |  |  |
| IIIb | 1.000 |  |  | 1.000 |  |  |
| IIIc | 1.449 | 0.947–2.215 | 0.0872 | 1.575 | 0.764–3.245 | 0.2185 |
| IV | 270.509 | 65.58–1115.82 | **<0.0001** | 3.501 | 1.869–6.559 | **<0.0001** |
| **Pathological Stage** |  |  |  |  |  |  |
| 0-1 | 1.000 |  |  | 1.000 |  |  |
| 2 | 2.962 | 1.216–7.215 | **0.0168** | 1.101 | 0.289–4.194 | 0.8875 |
| 3 | 4.155 | 2.149–8.031 | **<0.0001** | 1.880 | 0.746–4.735 | 0.1807 |
| 4 | 264.116 | 67.773–1029.27 | **<0.0001** | 2.784 | 1.057–7.336 | **0.0383** |
| **Estrogen receptor** |  |  |  |  |  |  |
| Negative | 1.000 |  |  | 1.000 |  |  |
| Positive | 0.830 | 0.630–1.095 | 0.1872 | 0.565 | 0.341–0.935 | **0.0263** |
| **Progesterone receptor** |  |  |  |  |  |  |
| Negative | 1.000 |  |  | 1.000 |  |  |
| Positive | 0.739 | 0.549–0.996 | **0.0469** | 0.501 | 0.279–0.901 | **0.0209** |
| **Hormone receptor** |  |  |  |  |  |  |
| Negative | 1.000 |  |  | 1.000 |  |  |
| Positive | 0.841 | 0.637–1.110 | 0.2211 | 0.630 | 0.381–1.041 | 0.0715 |
| **HER2** |  |  |  |  |  |  |
| Negative | 1.000 |  |  | 1.000 |  |  |
| Positive | 0.743 | 0.546–1.010 | 0.0582 | 0.663 | 0.377–1.165 | 0.1531 |
| **Hormone receptor / HER2** |  |  |  |  |  |  |
| HR+/HER2- | 1.000 |  |  | 1.000 |  |  |
| HR+/HER2+ | 0.745 | 0.469–1.181 | 0.2103 | 0.510 | 0.193–1.349 | 0.1749 |
| HR-/HER2+ | 0.845 | 0.562–1.271 | 0.4190 | 1.124 | 0.559–2.258 | 0.7429 |
| HR-/HER2- | 1.303 | 0.933–1.821 | 0.1204 | 1.604 | 0.857–3.003 | 0.1397 |
| **Grade** |  |  |  |  |  |  |
| I-II | 1.000 |  |  | 1.000 |  |  |
| III | 1.102 | 0.786–1.546 | 0.5720 | 0.928 | 0.500–1.725 | 0.8140 |
| **Lymphatic invasion<sup>1</sup>** |  |  |  |  |  |  |
| Negative | 1.000 |  |  | 1.000 |  |  |
| Positive | 1.698 | 1.201–2.402 | **0.0027** | 2.331 | 1.222–4.450 | **0.0103** |
| **Vascular invasion<sup>1</sup>** |  |  |  |  |  |  |
| Negative | 1.000 |  |  | 1.000 |  |  |
| Positive | 1.784 | 1.261–2.525 | **0.0011** | 2.419 | 1.266–4.619 | **0.0075** |
| **Adjuvant chemotherapy** |  |  |  |  |  |  |
| No | 1.000 |  |  | 1.000 |  |  |
| Yes | 0.394 | 0.262–0.592 | **<0.0001** | 0.443 | 0.195–1.009 | 0.0526 |
| **Adjuvant radiation** |  |  |  |  |  |  |
| No | 1.000 |  |  | 1.000 |  |  |
| Yes | 0.175 | 0.126–0.242 | **<0.0001** | 0.344 | 0.206–0.572 | **<0.0001** |

sHR: subdistribution hazard ratio; <sup>1</sup>These covariates were not considered in multivariable models due to too many missing measurements.

Patients with low levels of serum S100A8/A9 had significantly better OS (*P*=0.0107, Figure 1a) and BCSS (*P*=0.005, Figure 1b). The OS probability at 60 months for the entire group was 0.54 (95% confidence interval [CI] 0.48–0.61); for S100A8/A9-low, 0.58 (95% CI 0.50–0.65); and for S100A8/A9-high, 0.45 (95% CI 0.31–0.58). The 60-months BCSS probability for the entire group was 0.56 (95% CI 0.50–0.63); for S100A8/A9-low, 0.56 (95% CI 0.50–0.63); and for S100A8/A9-high, 0.46 (95% CI 0.32–0.59). Median survival times were also shorter in the S100A8/A9-high group (OS=53 months, BCSS=53 months) relative to the S100A8/A9-low group (OS=96 months; BCSS=112 months). Notably, S100A8/A9 expression was significantly associated with an increased cumulative incidence of brain metastases (*P*=0.0159, Figure 1d), but not with metastases at other sites (*P*=0.3486, Figure 1c). Furthermore, IBC patients with brain metastases had worse OS than did those without brain metastases (*P*<0.0001, Figure 2a); in relation to any metastases, significant difference in the outcome was also observed (*P*<0.0001, Suppl Figure S1a).

**Figure 1.**
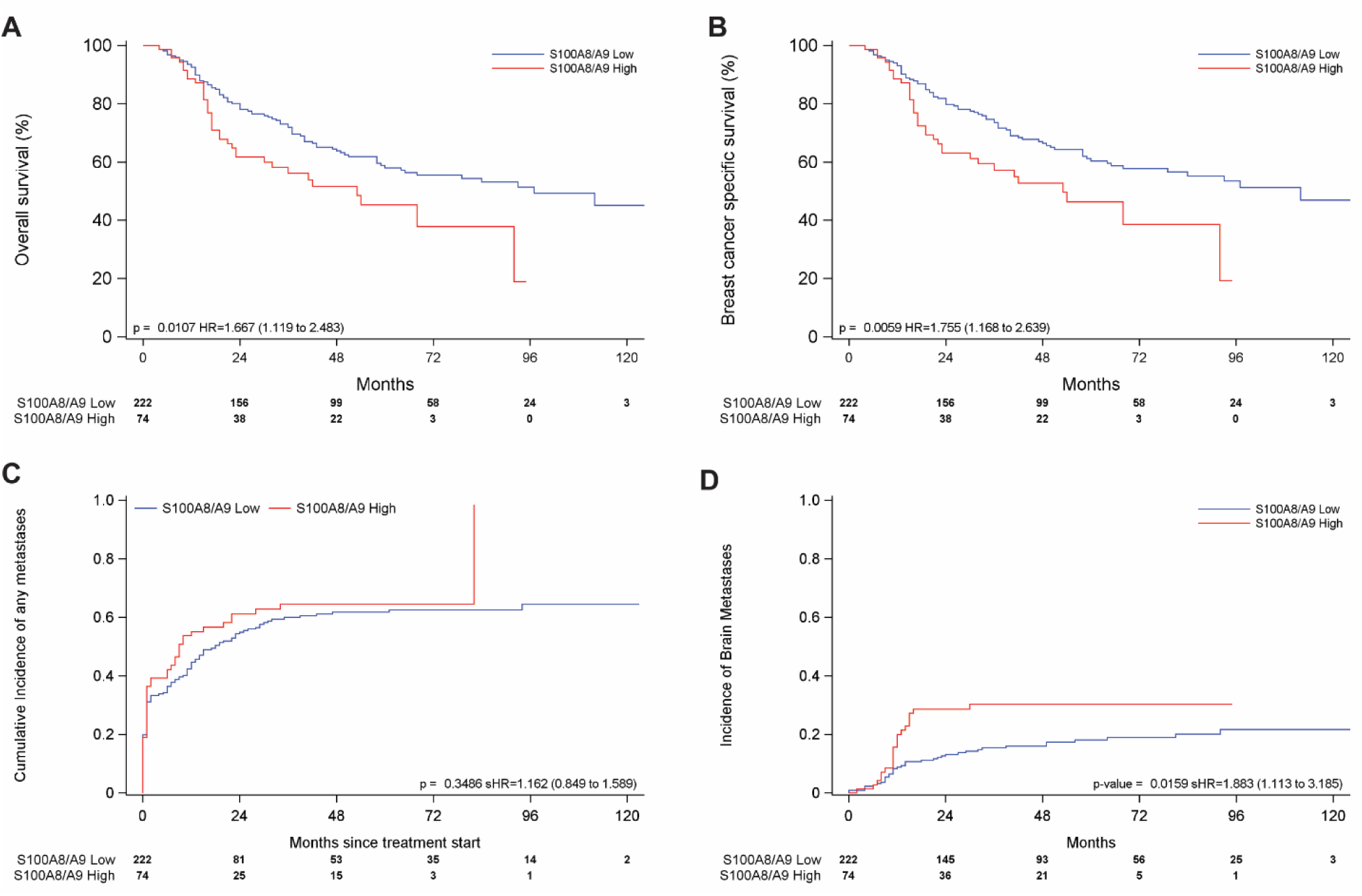
S100A8/A9 is associated with poor outcome in patients with IBC. Kaplan–Meier analysis for **(a)** overall survival and **(b)** breast cancer–specific survival by serum S100A8/A9 level. Incidence of **(c)** any metastases and **(d)** brain metastases by serum S100A8/A9 level. Abbreviations: HR = hazard ratio, sHR = subdistribution hazard ratio.

**Figure 2.**
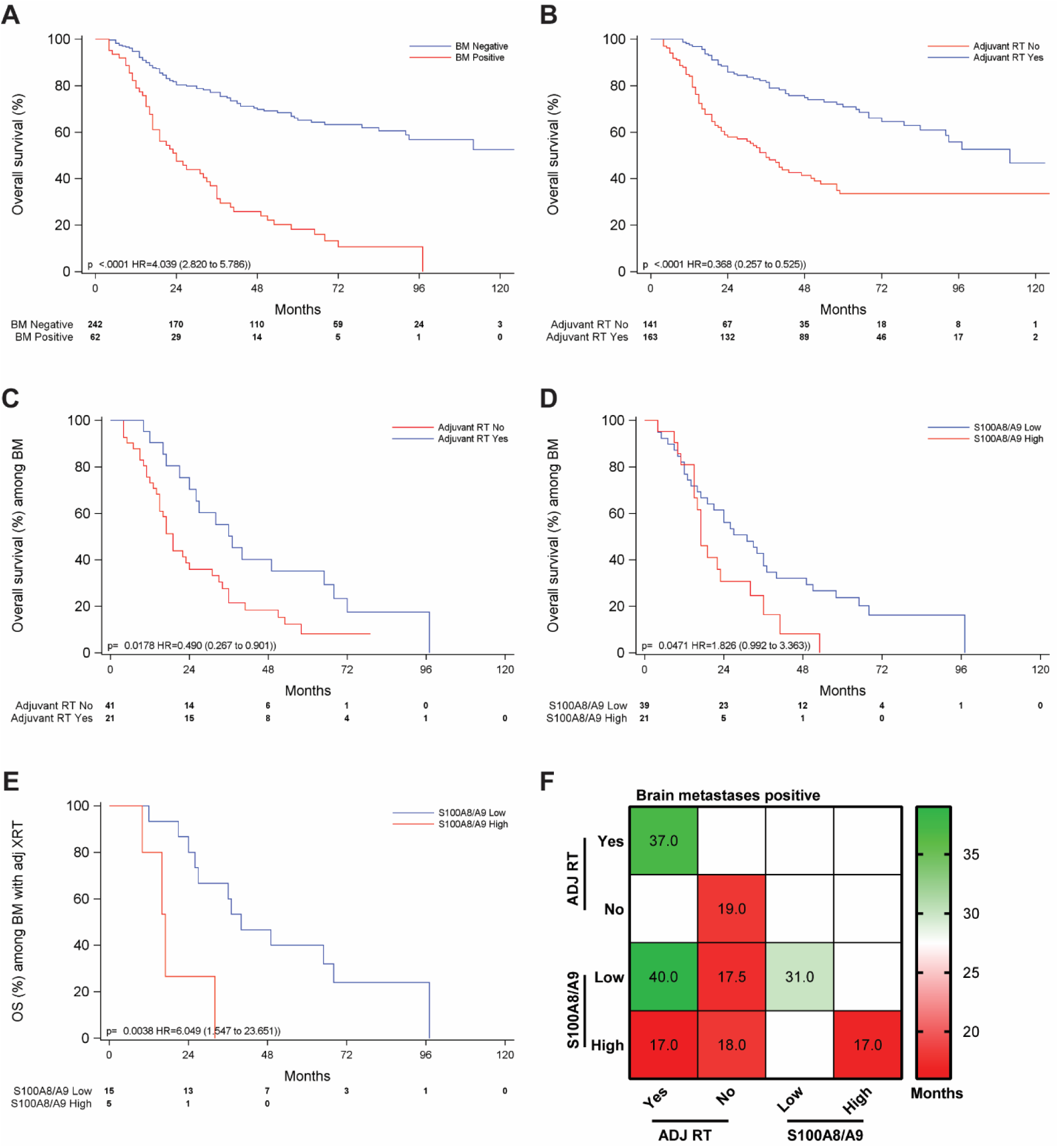
Kaplan–Meier analysis for **(a)** overall survival of IBC patients with (positive) or without (negative) brain metastases (BM), **(b)** overall survival (OS) by receipt or no receipt of adjuvant radiation (RT), **(c)** OS by receipt of adjuvant radiation in patients with brain metastases, **(d)** brain metastases OS by S100A8/A9 levels, **(e)** OS for patients with brain metastases and treated with adjuvant radiation (ADJ XRT), and **(f)** median overall survival times, in months, for patients positive for brain metastases, stratified by adjuvant radiation (ADJ RT) and S100A8/A9 levels.

In the multivariable analysis, independent predictors of OS and BCSS included S100A8/A9 levels, clinical disease stage, hormone receptor / HER2 status, and receipt of adjuvant radiation therapy (Table 4). Notably, high S100A8/A9 levels emerged as a strong and independent predictor of poorer survival outcomes, with an HR of 1.72 (95% CI 1.13–2.62; *P*=0.014) for OS and 1.83 (95% CI 1.19–2.82; *P*=0.0061) for BCSS. Regarding brain metastases, S100A8/A9 levels, clinical stage, and receipt of adjuvant radiation therapy were included in the multivariate analysis (Table 4). High levels of S100A8/A9 were independently and significantly associated with an increased incidence of brain metastases, with a sHR of 1.8 (95% CI 1.07–3.04; *P*=0.0265).

**Table 4.** Multivariate Cox regression analysis for overall survival, breast cancer-specific survival, and multivariable Fine-Gray model on development of brain metastases among patients with IBC.

| Covariate | Overall survival |  |  | Breast cancer specific survival |  |  | Development of brain metastases |  |  |
| --- | --- | --- | --- | --- | --- | --- | --- | --- | --- |
|  | HR | 95% CI | P Value | HR | 95% CI | P Value | sHR | 95% CI | P Value |
| S100A9 |  |  |  |  |  |  |  |  |  |
| Low | 1.000 |  |  | 1.000 |  |  | 1.000 |  |  |
| High | 1.722 | 1.130–2.623 | **0.0114** | 1.829 | 1.188–2.816 | **0.0061** | 1.804 | 1.071–3.037 | **0.0265** |
| Clinical stage |  |  |  |  |  |  |  |  |  |
| IIIb | 1.000 |  |  | 1.000 |  |  | 1.000 |  |  |
| IIIc | 2.240 | 1.352–3.713 | **0.0017** | 2.233 | 1.307–3.815 | **0.0033** | 1.457 | 0.693–3.063 | 0.3206 |
| IV | 1.169 | 0.633–2.159 | 0.6178 | 1.298 | 0.683–2.467 | 0.4266 | 1.885 | 0.869–4.092 | 0.1088 |
| Hormone receptor / HER2 |  |  |  |  |  |  |  |  |  |
| HR+/HER2- | 1.000 |  |  | 1.000 |  |  |  |  |  |
| HR+/HER2+ | 0.373 | 0.188–0.742 | **0.0049** | 0.357 | 0.173–0.735 | **0.0052** |  |  |  |
| HR-/HER2+ | 0.377 | 0.197–0.718 | **0.0030** | 0.399 | 0.208–0.765 | **0.0057** |  |  |  |
| HR-/HER2- | 2.103 | 1.384–3.194 | **0.0005** | 2.119 | 1.375–3.267 | **0.0007** |  |  |  |
| Adjuvant radiation |  |  |  |  |  |  |  |  |  |
| No | 1.000 |  |  | 1.000 |  |  | 1.000 |  |  |
| Yes | 0.307 | 0.178–0.531 | **<0.0001** | 0.301 | 0.169–0.538 | **<0.0001** | 0.448 | 0.218–0.918 | **0.0283** |
| Age |  |  |  |  |  |  |  |  |  |
| 1 Unit Change |  |  |  |  |  |  | 0.975 | 0.953–0.997 | **0.0288** |

Adjuvant radiation therapy also emerged as an independent variable with a strong association with OS (HR=0.31, 95% CI 0.18–0.53, *P*<0.0001), BCSS (HR=0.30, 95% CI 0.17–0.54, *P*<0.0001), and development of brain metastases (HR=0.45, 95% CI 0.22–0.92, *P=*0.0283) (Table 4). Patients who received adjuvant radiation exhibited significantly better OS than those who did not (OS, *P*<0.0001; Figure 2b). Within the radiation-treated cohort, patients with high serum S100A8/A9 levels may have had poorer outcomes than those with low S100A8/A9 levels (OS, *P*=0.081; Suppl Figure S1b). In the subgroup of patients not treated with adjuvant radiation, S100A8/A9 levels may have influenced survival outcomes (OS, *P*=0.065; Suppl Figure S1c). The median OS time for all patients who received adjuvant radiation was 112.0 months versus 36 months for those who did not receive radiation (Suppl Table S1). When stratified by S100A8/A9 levels, patients with S100A8/A9-high levels had a median survival time of 68 months, whereas the median survival time was not reached in the S100A8/A9-low group (Suppl Table S2), indicating a significant survival advantage for patients with low S100A8/A9 levels.

The presence of brain metastases was also associated with significantly reduced OS (Figure 2a). However, the addition of adjuvant radiation therapy was linked to improved outcomes in these patients (*P*=0.018; Figure 2c). Notably, patients with low serum levels of S100A8/A9 also exhibited better survival outcomes despite the presence of brain metastases (*P*=0.05; Figure 2d); while no difference in the outcome was observed in those with metastases at other sites (*P*=0.25, Suppl Figure S1b). Interestingly, within the subgroup of patients who received adjuvant radiation and had brain metastases, those with low S100A8/A9 levels demonstrated better survival than those with high S100A8/A9 levels (*P*=0.004; Figure 2e), with median survival times of 37 months *vs.* 19 months, respectively (Figure 2f, Suppl Tables S3 and S4); however, no significant difference was observed within the subgroup that had brain metastasis but did not receive adjuvant radiation (Suppl Figure S1D).

## Discussion

IBC remains a relatively under-characterized disease, lacking defined and specific therapeutic targets and prognostic biomarkers. In this study, we showed that patients with high S100A8/A9 serum levels had poorer OS and a higher likelihood of developing brain metastases. Multivariate analysis confirmed these levels as an independent predictor of reduced survival and increased brain metastatic risk. These findings emphasize that S100A8/A9 serum levels may serve as a valuable biological marker for stratifying IBC patients to personalize surveillance and therapeutic strategies.

Elevated S100A8/A9 expression has been previously implicated in tumor progression, invasion, and metastasis through activation of pattern recognition receptors such as TLR4 and RAGE, thereby promoting pro-inflammatory signaling and immune evasion^13-15^. S100A8 and S100A9 are key mediators in establishing pre-metastatic niches in both the brain and lung, creating a supportive microenvironment that facilitates tumor cell colonization and survival. In the brain metastatic niche, S100A9 has been shown to drive therapy resistance. Monteiro et al. demonstrated that S100A9 promotes radioresistance and reduces the efficacy of whole-brain irradiation^20^. Complementing this, Biswas et al. reported that in EGFR-mutant lung cancer, S100A9 enhances brain metastatic outgrowth and relapse, and that targeting S100A9 or associated mechanisms impairs metastatic progression^19^. Other studies have also demonstrated elevated S100A9 levels to be linked with disease progression, poorer survival outcomes, and resistance to immunotherapies^20,25,26^. Together, these studies highlight the central role of S100A9, and the S100A8/A9 complex, in shaping brain and lung metastatic niches, promoting radioresistance, and driving metastatic relapses, making them attractive targets for therapeutic intervention and biomarker-guided strategies. Consistent with these preclinical and clinical observations, we found that high S100A8/S100A9 serum levels correlated with increased death and brain metastases in IBC patients. Given S100A8/A9’s established role in shaping pre-metastatic niches through immune modulation and inflammation, it may have broader relevance in other aggressive cancer subtypes prone to brain dissemination.

Adjuvant radiation therapy is a critical component of breast cancer management, particularly for IBC, and known to improve disease-specific survival and reduce the risk of tumor relapse^30-32^. In our cohort, adjuvant radiation emerged as a strong independent predictor of improved outcomes across several endpoints, including OS, BCSS, and decreased incidence of brain metastases. Specifically, patients who received adjuvant radiation had a median overall survival time of 111 months compared with 36 months for those who did not, underscoring its significant clinical benefit. This observation may be influenced by baseline patient characteristics, as poorer performance status and comorbidities could both limit eligibility for radiation therapy and contribute to inferior outcomes. Interestingly, among patients treated with radiation, those with high serum S100A8/A9 levels may have had worse survival relative to patients with low levels, suggesting that elevated S100A8/A9 may reduce the therapeutic benefit of radiation treatment. These observations suggest that the negative impact of elevated S100A8/A9 on survival may be mechanistically driven by their ability to promote radioresistance and metastatic relapse through pathways such as S100A9–RAGE–NF-κB–JunB signaling and ALDH1A1–retinoic acid activation, as demonstrated in preclinical models of brain metastases^19,20^. Recent evidence supports this notion: Monteiro et al. demonstrated that activation of the S100A9–RAGE–NF-κB–JunB axis drives resistance to whole-brain radiotherapy by enhancing pro-survival programs and reducing radiation-induced apoptosis. Elevated S100A9 levels, detected both within metastatic lesions and in patient plasma, correlated with poor radiotherapy response, supporting its utility as a predictive biomarker for stratifying patients undergoing brain-directed radiation^20^. Further, Biswas et al. found that in EGFR-mutant lung cancer brain metastases, high intracellular S100A9 cooperates with upregulation of ALDH1A1 and activation of the retinoic acid signaling pathway to drive lethal brain relapses, and that suppression of S100A9, ALDH1A1, or retinoic acid receptors markedly diminished brain metastatic growth^19^. Although S100A8 has been less directly implicated in the radioresponse of brain metastases, its established anti-apoptotic role after irradiation and its capacity to function as a heterodimeric partner with S100A9 suggest a broader involvement of the S100A8/A9 complex in modulating treatment resistance and the metastatic microenvironment^13,33^. Collectively, these studies position S100A9, and potentially the S100A8/A9 complex, as compelling therapeutic targets to overcome radioresistance and brain relapse in metastatic disease, providing a strong rationale for confirmatory studies and integrating S100A8/A9-directed inhibitors or biomarker-guided patient selection strategies into future neuro-oncology protocols.

In summary, we show that elevated serum level of S100A8/A9 is an independent prognostic indicator of poor outcome and increased brain metastasis risk in patients with IBC. These findings highlight the potential of S100A8/A9 as a predictive biomarker for identifying patients at high risk of developing brain metastasis in patients with IBC and other aggressive cancer subtypes.

## Methods

### Patient Serum Samples

This study was approved by the Institutional Review Board of The University of Texas MD Anderson Cancer Center (IBC Registry protocol 2006-1072; LAB Protocol – PA15-0802). Written informed consent was obtained from all patients before study enrollment. This study is compliant with all relevant ethical regulations on the use of human serum, and all research was performed in accordance with the Declaration of Helsinki. Serum samples were obtained from all participants at study initiation or, when this was not possible, at the earliest feasible subsequent time point. Samples were drawn from several cohorts and included patients who were either newly diagnosed or previously treated. Patient characteristics are described in Table 1.

### ELISA

A Human S100A8/S100A9 Heterodimer ELISA Kit - Quantikine (DS8900; R&D Systems, Minneapolis, MN, USA) was used to measure the serum levels of S100A8/A9 in 304 patients according to the manufacturer’s instructions.

### Statistical Analysis

The cutoff for S100A8/A9 levels was the highest quadrant (Q3), placing 222 patients as S100A8/A9-low (≤5108 ng/mL) and 74 as S100A8/A9-high (>5108 ng/mL). Patient characteristics were summarized by S100A8/A9 value (low [≤5108 ng/mL] vs. high [>5108 ng/mL]) and compared between patients with S100A8/A9-low and S100A8/A9-high serum levels. Univariate and multivariate Cox regression analyses were performed considering overall survival and breast cancer–specific survival as outcome variables. Univariate and multivariate Fine-Gray models were performed considering time to develop metastasis and time to develop brain metastasis as outcome variables. Death without an event of interest was considered as a competing risk event in the Fine-Gray model. The Kaplan-Meier method was used to estimate survival probability, and cumulative incidence was estimated by Aalen-Johansen method. Log-rank tests were used to compare survival curves. The Gray test was used to compare cumulative incidence plots. *P* values of < 0.05 were taken to indicate statistical significance. SAS 9.4 (SAS institute INC, Cary, NC) was used for data analysis.

## Data Availability

The datasets generated and/or analyzed during the current study are not publicly available due to the confidential nature of the clinical data but are available from the corresponding author in patient ID-redacted form upon reasonable request.

## Acknowledgements

We thank Christine F. Wogan, MS, ELS, of MD Anderson’s Division of Radiation Oncology for scientific review and editing of the manuscript.

## Funding

This work was supported by grants from American Cancer Society Research Scholar grant (RSG-19-126-01 and ACS-TLC-23-997857-01 to B.G.D.), MD Anderson Cancer Center (Bridge Funding, Startup, and Bootwalk funds), the State of Texas Grant for Rare and Aggressive Cancers, and the National Cancer Institute Cancer Center Support (Core) Grant P30 CA016672 to The University of Texas MD Anderson Cancer Center (PI: PW Pisters). The statistical analysis work was supported in part by the Cancer Center Support Grant (NCI Grant P30 CA016672).

## Author contributions

E.S.V. conceived and designed the project, performed experiments, analyzed data and interpreted the results, wrote and edited the manuscript with input from all other authors. X.H. performed experiments, analyzed data and interpreted the results. J.S. provided statistical analysis support. K.G. performed experiments. E.N.C., J.M.R, A.N., B.L., D.T., W.A.W, and S.K. provided resources and contributed to the revision of the manuscript. B.G.D. conceived and designed the project, supervised the study, interpreted the results, wrote and edited the manuscript with input from all other authors. All authors have read and agreed to the published version of the manuscript.

## Contributors: MDACC Inflammatory Breast Cancer Team Authorship

The prospective IBC annotated biobank exists through the collaborative efforts of the following individuals who manage the protocol and regulatory compliance, meet regularly to review and adjudicate inconclusive diagnoses, accrue and consent patients, coordinate sample collections, maintain records, review data collection and specimen inventory, and provide researchers access to the materials through secondary IRB approved protocols: Rachel Layman, Bora Lim, Azadeh Nasrazadani, Sadia Saleem, Vicente Valero, Michael C. Stauder, Wendy A. Woodward, Anthony Lucci, Susie X. Sun, Gary J. Whitman, Miral Patel, Huong Le-Petross, Yang Lu, Angela Marx, Angela Alexander, Chasity Yajima, Megumi Kai, Lily Villareal, and Heather Lopez.

## Competing interests

The authors declare no competing interests.

## Additional information

Supplementary information available online

## Notes

### Competing Interest Statement

The authors have declared no competing interest.

### Author Declarations

This study was approved by the Institutional Review Board of The University of Texas MD Anderson Cancer Center (IBC Registry protocol 2006-1072; LAB Protocol PA15-0802).

