## Supplementary Figure and Tables for "Serum S100A8/S100A9 is associated with increased risk of brain metastasis in patients with inflammatory breast cancer"

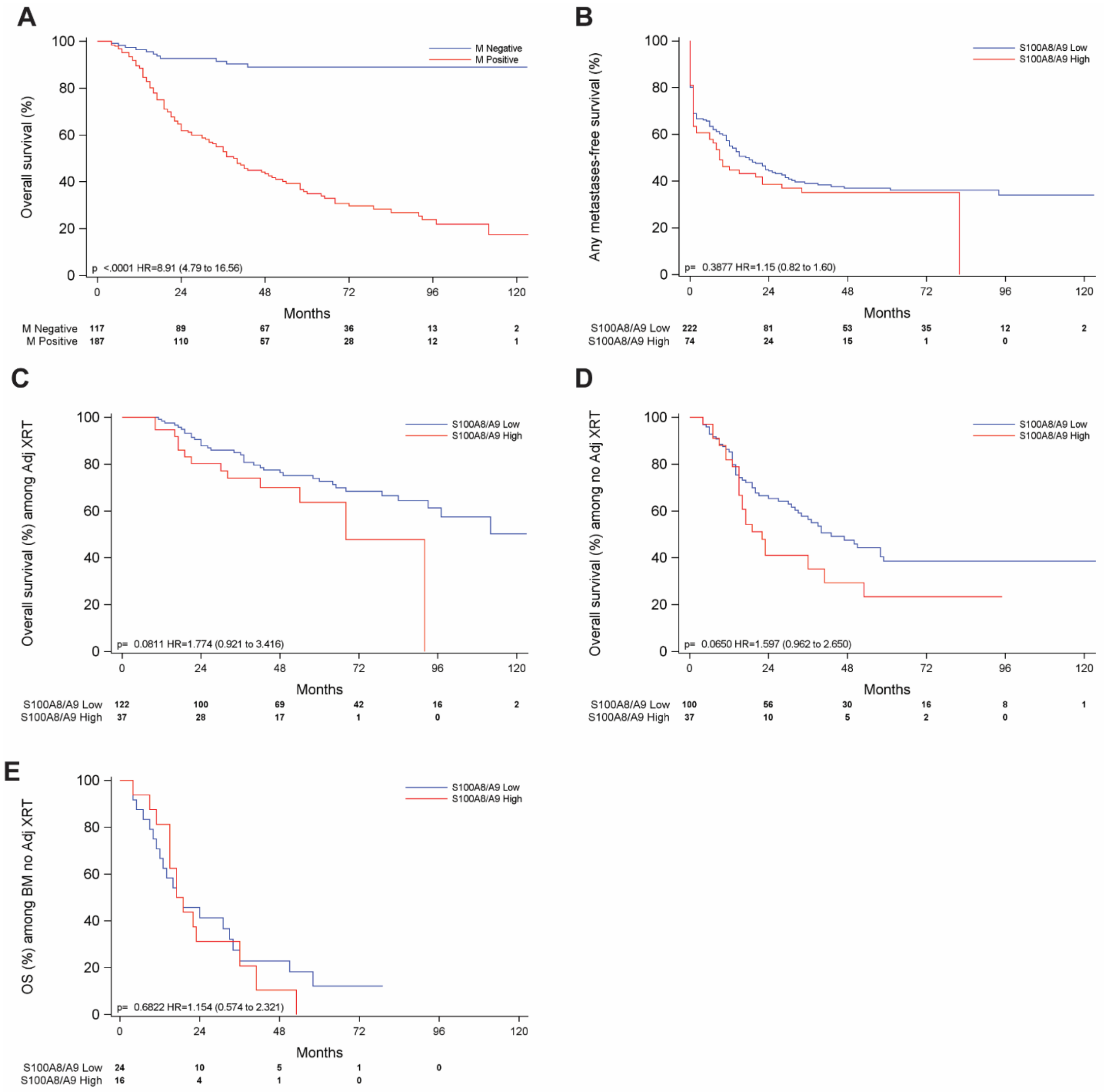


**Supplementary Figure 1.** Kaplan–Meier analysis for **(a)** any metastases overall survival **(b)** any metastasis according to S100A8/A9 levels, **(c)** overall survival for patients who received adjuvant radiation (ADJ XRT) according to S100A8/A9 levels, **(d)** overall survival for patients who did not receive adjuvant radiation according to S100A8/A9 levels, and **(e)** overall survival for patients who did not receive adjuvant radiation and had brain metastases, by S100A8/A9 levels.

**Supplementary Table S1:** Median and 95% confidence interval [CI] patients' survival (months)

| Covariate | Median | 95% CI |
|---|---|---|
| **Brain metastases** | | |
| Positive | 24.0 | 17.0–34.0 |
| Negative | Undefined | N/A |
| **Adjuvant radiation** | | |
| Yes | 112.0 | 92.0–+ infinity |
| No | 36.0 | 23.0–50.0 |
| **S100A8/A9** | | |
| Low | 97.0 | 64.0–+ infinity |
| High | 53.0 | 23.0–+ infinity |

**Supplementary Table S2:** Median and 95% confidence interval [CI] patients' survival stratified by adjuvant radiation and S100A8/A9 levels (months)

| Covariate | Median | 95% CI |
|---|---|---|
| **Adjuvant radiation Yes** | | |
| S100A8/A9 Low | Undefined | N/A |
| S100A8/A9 High | 68.0 | 54.0–92 |
| **Adjuvant radiation No** | | |
| S100A8/A9 Low | 43.0 | 32.0–59.0 |
| S100A8/A9 High | 22.0 | 15.0–40.0 |

**Supplementary Table S3:** Median and 95% confidence interval [CI] patients' survival with brain metastases positive (months)

| Covariate | Level | Median | 95% CI |
|---|---|---|---|
| **Adjuvant radiation** | | | |
| Yes | | 37.0 | 24.0–65.0 |
| No | | 19.0 | 15.0–31.0 |
| **S100A8/A9** | | | |
| Low | | 31.0 | 17.0–37.0 |
| High | | 17.0 | 15.0–32.0 |

**Supplementary Table S4:** Median patients' survival with brain metastases positive and received adjuvant radiation (months)

| Covariate | Median survival | 95% confidence interval |
|---|---|---|
| **S100A8/A9** | | |
| Low | 40.0 | 24.0–68.0 |
| High | 17.0 | 10.0–32.0 |
